# Using deep learning to analyze the compositeness of musculoskeletal aging reveals that spine, hip and knee age at different rates, and are associated with different genetic and non-genetic factors

**DOI:** 10.1101/2021.06.14.21258896

**Authors:** Alan Le Goallec, Samuel Diai, Sasha Collin, Théo Vincent, Chirag J. Patel

**Affiliations:** Department of Biomedical Informatics, Harvard Medical School, Boston, MA, 02115, USA; Department of Systems, Synthetic and Quantitative Biology, Harvard University, Cambridge, MA, 02118, USA

## Abstract

With age, the musculoskeletal system undergoes significant changes, leading to diseases such as arthritis and osteoporosis. Due to the aging of the world population, the prevalence of such diseases is therefore expected to starkly increase in the coming decades. While numerous biological age predictors have been developed to assess musculoskeletal aging, it remains unclear whether these different approaches and data capture a single aging process, or if the diverse joints and bones in the body age at different rates. In the following, we leverage 42,000 full body, spine, hip and knee X-ray images and musculoskeletal biomarkers from the UK Biobank and use artificial intelligence to build the most accurate musculoskeletal aging predictor to date (RMSE=2.65±0.01 years; R-Squared=87.6±0.1%). Our predictor is composite and can be used to assess spine age, hip age and knee age, in addition to general musculoskeletal aging. We find that accelerated musculoskeletal aging is moderately correlated between these different musculoskeletal dimensions (e.g hip vs. knee: Pearson correlation=.351±.004). Musculoskeletal aging is heritable at more than 35%, and the genetic factors are partially shared between joints (e.g hip vs. knee: genetic correlation=.52±.04). We identified single nucleotide polymorphisms associated with accelerated musculoskeletal aging in approximately ten genes for each musculoskeletal dimension. General musculoskeletal aging is for example associated with a TBX15 variant linked to Cousin syndrome and acromegaloid facial appearance syndrome. Finally, we identified biomarkers, clinical phenotypes, diseases, environmental and socioeconomic variables associated with accelerated musculoskeletal aging in each dimension. We conclude that, while the aging of the different components of the musculoskeletal system is connected, each bone and joint can age at significantly different rates.

## Background

With age, the musculoskeletal system undergoes important changes, putting individuals at risk for diseases ^1, 2^. Bone shape, density and cell composition are affected, leading to osteoporosis and an increased risk of fracture ^3–5^. Aging affects the joints, increasing the risk for osteoarthritis ^6^ and degeneration of the intervertebral discs ^7^, as well as muscle mass and composition (sarcopenia) ^8^. These changes lead to a greater vulnerability to falls, resulting in increased mortality, morbidity and health care cost. 28.7% of US elderly fell in 2014 ^9^.

Biological age predictors have been built to better understand the aging process of the musculoskeletal system. In contrast to chronological age --a mere measure of the time that passed since an individual’s birth-- biological age is a measure of the condition and damage afflicting the body, and is the true underlying cause of age-related diseases. A forty year-old patient suffering from early-onset osteoporosis and osteoarthritis could for example have a biological musculoskeletal age of fifty years: we refer to such an individual as an accelerated ager. Biological age predictors can be built by training machine learning algorithms to predict chronological age from biomedical features, with the biological age of a participant being defined as the prediction outputted by the model. Others have used full body ^10^, chest ^11^, hip ^12–15^, knee ^16–22^ or hand ^23–30^ X-ray images to predict age.

What remains elusive questions is whether the different bones, joints and muscle of a given individual age at the same rate, whether accelerated musculoskeletal aging is heritable, and what are the genetic factors associated with it. In the following, we leverage 42,000 UK Biobank ^31^ full body, spine, hip and knee X-ray images, along with musculoskeletal biomarkers from 37-82 year-old participants,and use deep learning to build biological age predictors capturing different facets of musculoskeletal aging. We perform genome wide association studies [GWASs] and X-wide association studies [XWAS] to identify genetic and non-genetic factors (e.g diseases, environmental and socioeconomic variables) associated with accelerated aging in these different musculoskeletal dimensions.

## Results

### We predicted chronological age within three years

We leveraged the UK Biobank, a dataset containing 42,000 full body, spine (sagittal and coronal views), hip and knee X-ray images (Figure 1A, B and C), as well as more 500,000 than anthropometry, impedance, heel bone densitometry and hand grip strengths measurements (Figure 1A and B) from patients aged 37-82 years (Fig. S1). We predicted chronological age from X-ray images using convolutional neural networks and from scalar biomarkers using elastic nets, gradient boosted machines [GBMs] and shallow fully connected neural networks. We then hierarchically ensembled these 31 models by musculoskeletal dimensions and subdimensions (Figure 1A and D).

**Figure 1:**
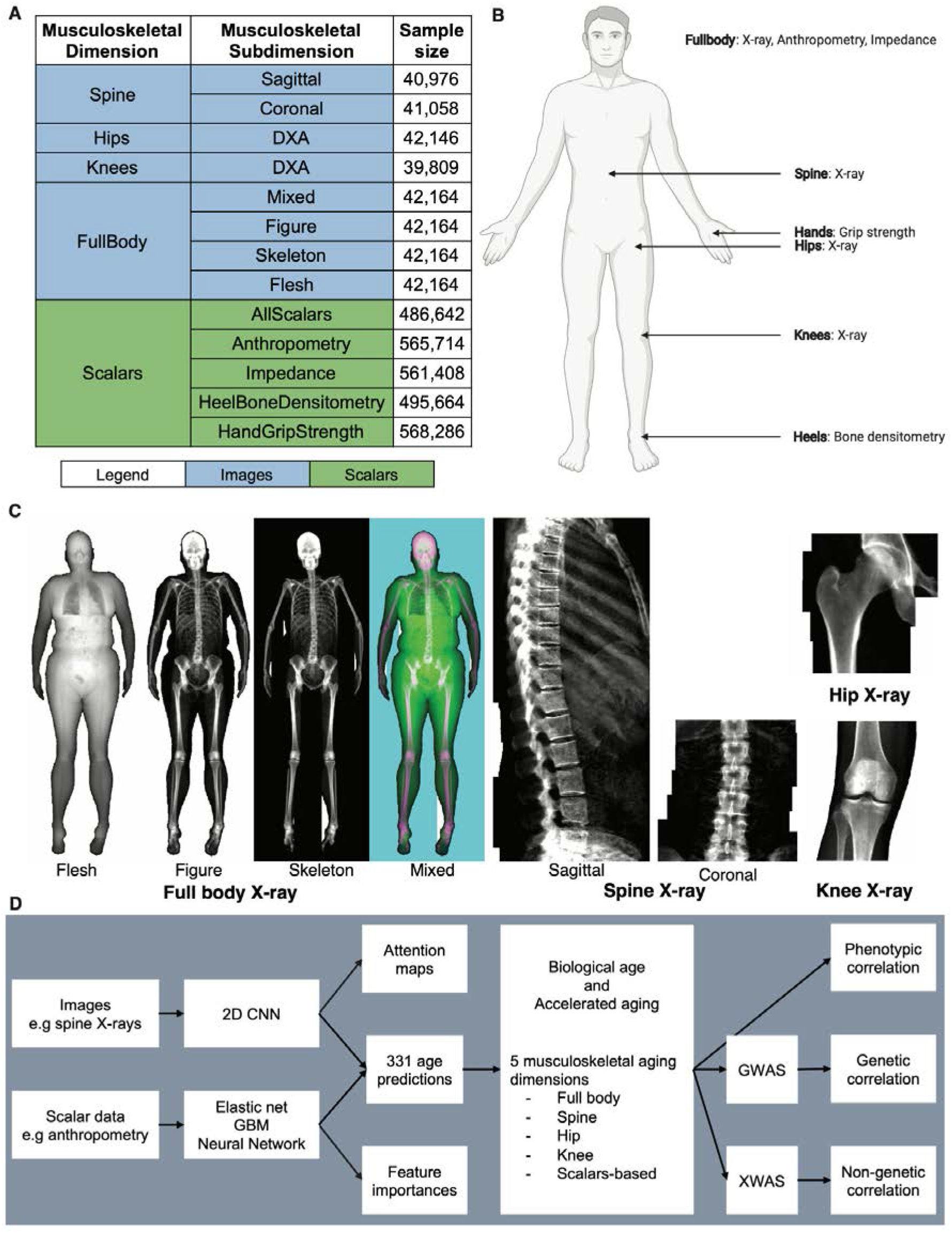
Overview of the datasets and analytic pipeline. A - Musculoskeletal aging dimensions and subdimensions. B - Body locations from where the datasets were collected. C - X-ray samples. D - Analytic pipeline.

We predicted chronological age with a testing root mean square error [RMSE] of 2.65±0.01 years and a R-Squared [R^2^] value of 87.6±0.1% (Figure 2). This prediction accuracy was largely driven by the full body X-ray-based model (R^2^=85.7±0.1%). The spine X-ray-based model was the second highest most accurate (R^2^=74.6±0.2%), with the sagittal view images providing more information (R^2^=71.3±0.2%) than the coronal view images (R^2^=64.5±0.3%). Hip and knee X-ray-based models performed the same (R^2^=69.0±0.3%), and the ensemble model built on scalar datasets (anthropometry, impedance, heel bone densitometry and hand grip strength) performed the worst (R^2^=25.9±0.1%), despite a 12 times larger sample size.

**Figure 2:**
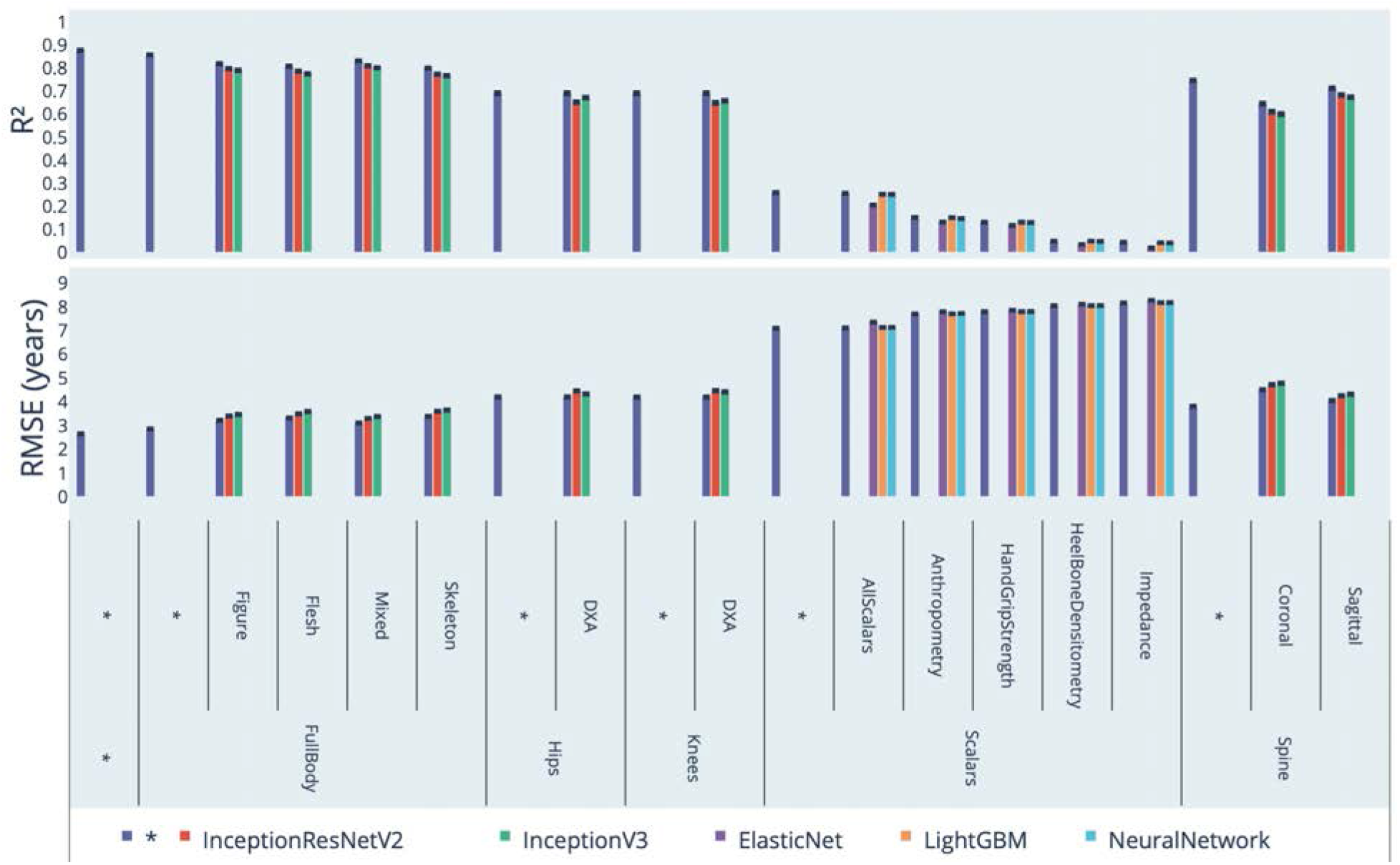
Chronological age prediction performance (R^2^ and RMSE) * represent ensemble models

### Identification of features driving musculoskeletal age prediction

We used attention maps to identify the features driving the prediction of image-based models (Figure 3).

**Figure 3:**
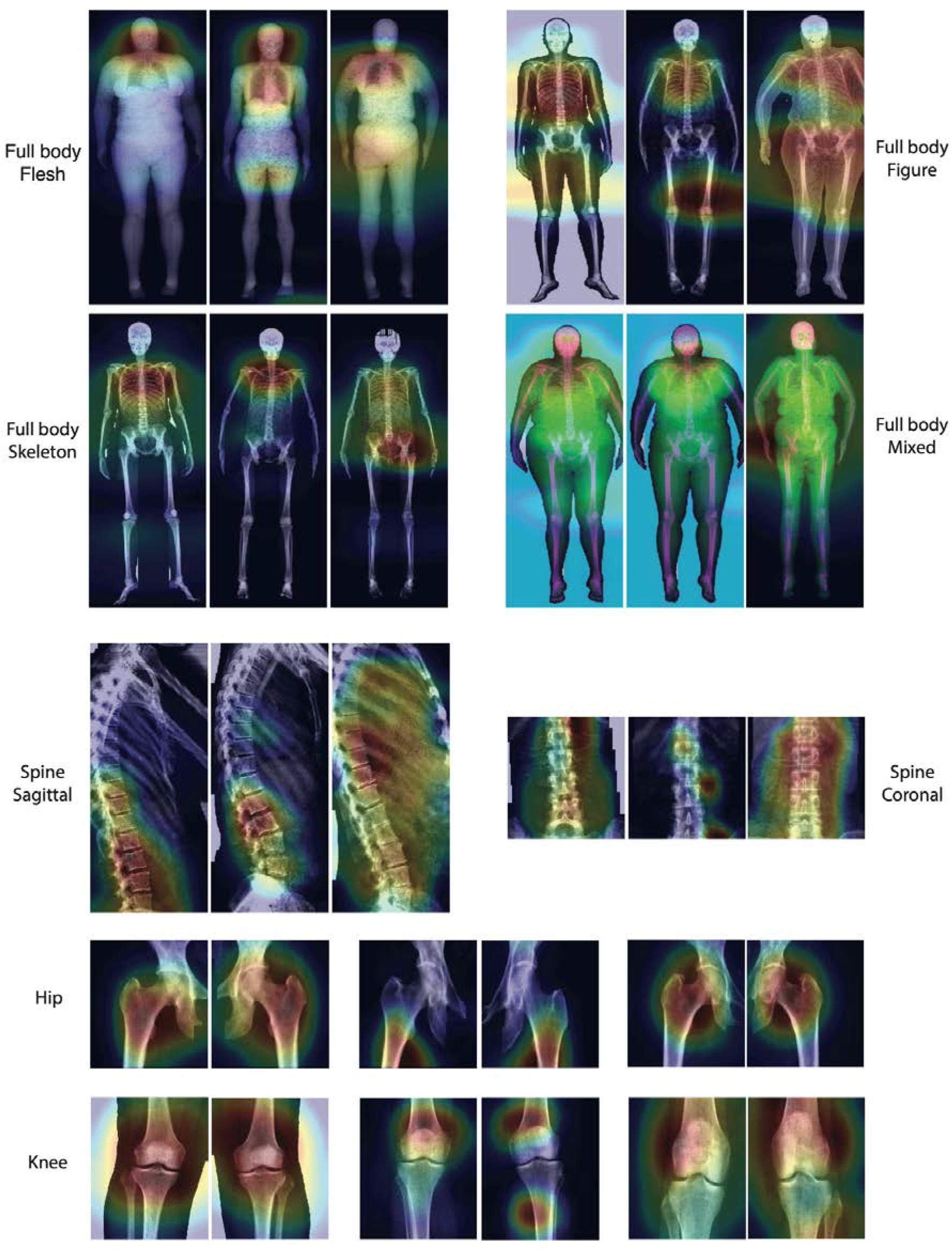
Attention map samples for X-ray-based models.

Warm colors highlight regions of high importance according to the Grad-RAM map. All images were collected from 60-65 year-old females. For each triplet of images, the leftmost image is a decelerated ager, the central image is a normal ager, and the rightmost image is an accelerated ager.

Attention maps for full body X-ray images highlighted diverse body parts across participants. Regions frequently highlighted included the neck, the upper torso, the hips and the knees (Figure 4). Attention maps for sagittal view spine X-ray images most commonly highlighted the lumbar region (Supplementary Figure 22), whereas the coronal view images highlighted diverse vertebras across individuals (Supplementary Figure 23). Attention maps for hip X-rays consistently highlighted the greater trochanter of the femur as well as the joint (Supplementary Figure 24). Attention maps for knee X-rays highlighted diverse regions across participants, such as the femur, the shin, the joint itself and occasionally the fibula (Supplementary Figure 25).

**Figure 4:**
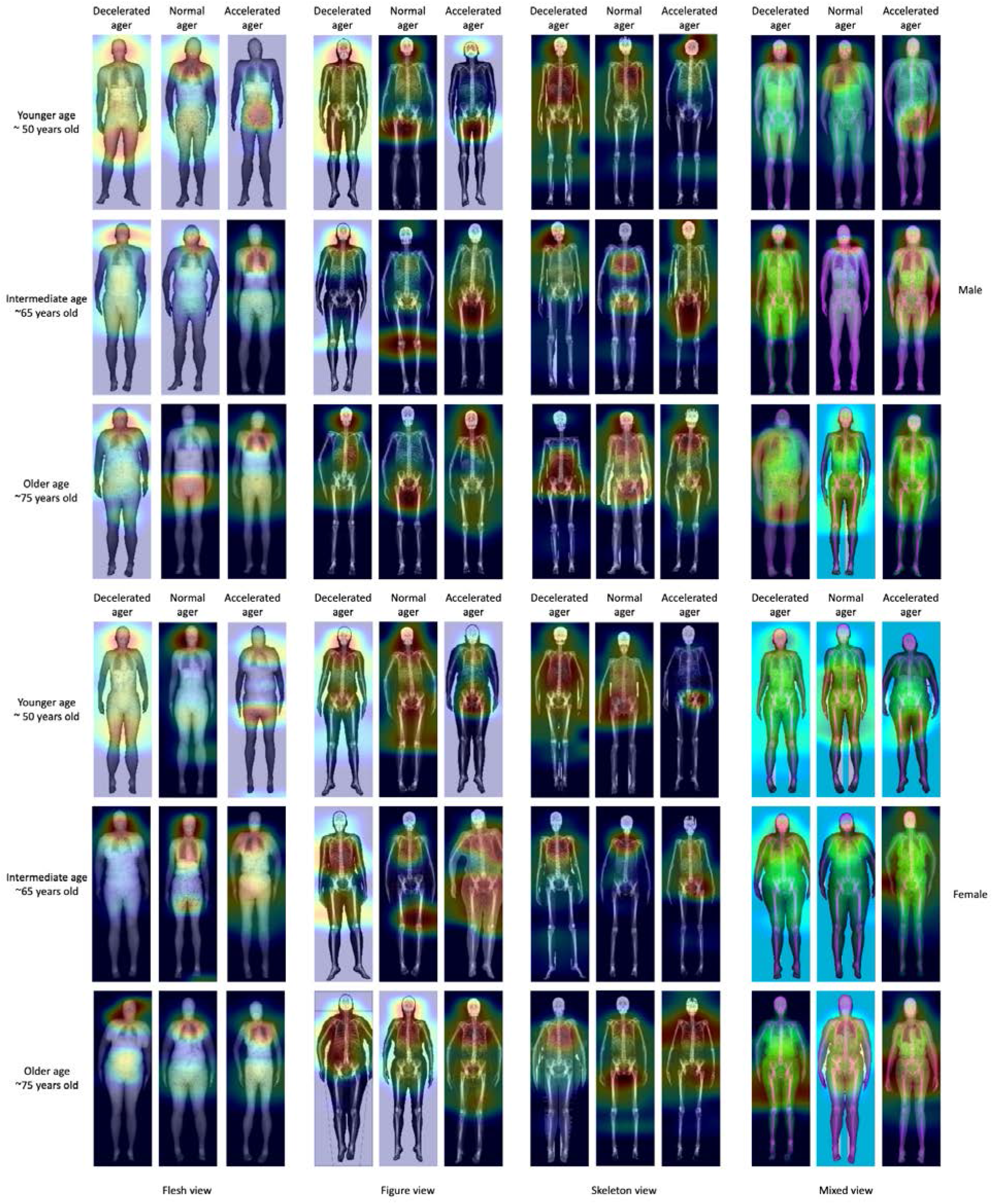
Attention maps for full body X-ray images. Warm colors highlight regions of high importance according to the Grad-RAM map.

**Figure 5:**
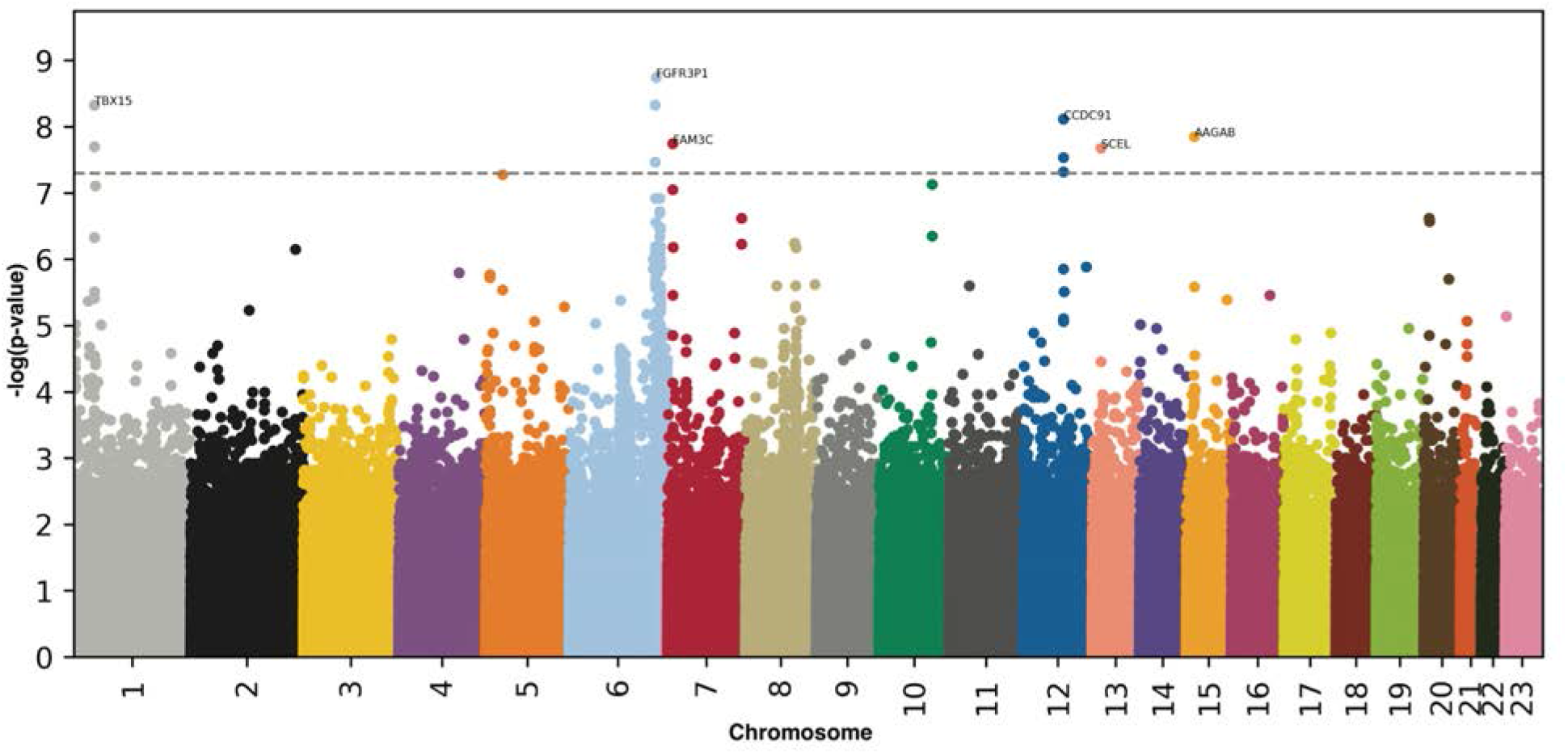
GWAS results - General musculoskeletal aging. -log10(p-value) vs. chromosomal position of locus. Dotted line denotes 5x10^-8^.

For scalar features, we best predicted accelerated musculoskeletal aging using a GBM or a neural network; however, the R^2^ was moderate (25.2±0.4%). Impedance measures (five features) and heel bone densitometry (six features) were poor predictors of chronological age, with respective R^2^ values of 4.2±0.2% and 4.9±0.2%. Prediction accuracy was driven by anthropometric measures (R^2^=15.1±0.3%; eight features) and hand grip strength (R^2^=13.1±0.3%; two features). Specifically, the most important features were (1) White ethnicity, (2) waist circumference, (3) weight, (4) impedance of whole body, (5) British ethnicity, (6) body mass index [BMI], (7) left hand grip strength, (8) Sex, (9) right arm impedance and (10) Indian ethnicity. The elastic net (R^2^=20.5±0.4%) assigned a positive regression coefficient to White ethnicity, waist circumference, and British ethnicity and a negative regression coefficient to weight, impedance of whole body, BMI, left hand grip strength and impedance of right arm.

### Genetic factors and heritability of accelerated musculoskeletal aging

We defined musculoskeletal age as the prediction outputted by the best performing model, after correction for the analytical bias in the residuals (see Methods). For example, spine age is defined as the prediction outputted by the ensemble model trained on both sagittal and coronal-view spine X-ray images, and accelerated spine aging is defined as the difference between spine age and chronological age.

We performed six genome wide association studies [GWASs] to estimate the GWAS-based heritability of general (h_g^2^=34.9±1.8%), full body X-ray-based (h_g^2^=30.7±1.6%), spine X-ray-based (h_g^2^=32.9±1.7%), hip X-ray-based (h_g^2^=27.7±1.6%), knee X-ray-based (h_g^2^=25.3±1.7%) and scalar biomarkers-based accelerated musculoskeletal aging (h_g^2^=22.1±0.1%). We identified 8-20 single nucleotide polymorphisms [SNPs] in 6-13 genes associated with accelerated aging for each X-ray-based aging dimension, and a larger number associated with accelerated aging as defined by the scalar-based ensemble model, due to the ∼10-times larger sample size. (Table 1)

**Table 1:** GWASs summary - Heritability, number of SNPs and genes associated with accelerated aging in each musculoskeletal dimension

| Musculoskeletal aging dimension | Sample size | SNPs | Genes | Heritability (%) | R-Squared for CA prediction (%) |
| --- | --- | --- | --- | --- | --- |
| General | 35,277 | 11 | 9 | 34.9±1.8 | 87.6±0.1 |
| Full body | 38,820 | 8 | 8 | 30.7±1.6 | 85.7±0.1 |
| Spine | 37,826 | 22 | 13 | 32.9±1.7 | 74.6±0.2 |
| Hip | 38,806 | 20 | 7 | 27.7±1.6 | 69.0±0.3 |
| Knee | 36,727 | 12 | 6 | 25.3±1.7 | 69.0±0.3 |
| Scalars-based | 407,998 | 2631 | 954 | 22.1±0.2 | 25.9±0.1 |

Approximately half (17/37) the peaks highlighted by the GWAS (e.g Figure 5) include a gene whose function can be linked to musculoskeletal function. Specifically, for accelerated general musculoskeletal aging, the GWAS highlighted TBX15 (linked to Cousin syndrome and acromegaloid facial appearance syndrome) and CCDC91 (associated with ossification of the posterior longitudinal ligament of the spine and diffuse idiopathic skeletal hyperostosis). For accelerated full body musculoskeletal aging, the GWAS highlighted TBX15 (linked to Cousin syndrome and acromegaloid facial appearance syndrome) and FGFR1 (linked to osteoglophonic dysplasia and Hartsfield syndrome). For accelerated spine aging, the GWAS highlighted TNFSF11 (linked to osteopetrosis), SUPT3H (linked to dysostosis), SOX5 (involved in cartilage formation and linked to Lamb-Shaffer syndrome) and ESR1 (linked to osteoporosis). For accelerated hip aging, the GWAS highlighted SUPT3H (linked to dysostosis), CCDC91 (associated with ossification of the posterior longitudinal ligament of the spine and diffuse idiopathic skeletal hyperostosis) and TNFSF11 (linked to osteopetrosis). For accelerated knee aging, the GWAS highlighted CCDC91 (associated with ossification of the posterior longitudinal ligament of the spine and diffuse idiopathic skeletal hyperostosis). For accelerated musculoskeletal scalar features-based (anthropometry, impedance, heel bone densitometry and hand grip strength) aging, the GWAS highlighted GDF5 (linked to osteoarthritis, acromesomelic dysplasia, brachydactyly, proximal symphalangism, chondrodysplasia and multiple synostoses syndrome), GPR126 (involved in body height and linked to distal arthrogryposis), PKDCC (involved in development and linked to rhizomelic limb shortening with dysmorphic features), POLD3 (linked to Ruijs-Aalfs syndrome) and ZBTB38 (involved in height).

We summarize our findings for each musculoskeletal dimension in the Supplementary.

### Biomarkers, clinical phenotypes, diseases, environmental and socioeconomic variables associated with accelerated musculoskeletal aging

We use “X” to refer to all nongenetic variables measured in the UK Biobank (biomarkers, clinical phenotypes, diseases, family history, environmental and socioeconomic variables). We performed an X-Wide Association Study [XWAS] to identify which of the 4,372 biomarkers classified in 21 subcategories (Table S5), 187 clinical phenotypes classified in 11 subcategories (Table S8), 2,073 diseases classified in 26 subcategories (Table S11), 92 family history variables (Table S14), 265 environmental variables classified in nine categories (Table S17), and 91 socioeconomic variables classified in five categories (Table S20) are associated (p-value threshold of 0.05 and Bonferroni correction) with accelerated musculoskeletal aging in the different dimensions. We summarize our findings for general accelerated musculoskeletal aging below. Please refer to the supplementary tables (Table S6, Table S7, Table S9, Table S10, Table S12, Table S13, Table S18, Table S19, Table S21, Table S22) for a summary of non-genetic factors associated with general, full body, spine, hip, knee, and scalar biomarkers-based accelerated musculoskeletal aging. The full results can be exhaustively explored at https://www.multidimensionality-of-aging.net/xwas/univariate_associations.

### Biomarkers associated with accelerated musculoskeletal aging

The three biomarker categories most associated with accelerated musculoskeletal aging are body impedance, pulse wave measurements and brain MRI weighted means. Specifically, 100.0% of anthropometry biomarkers are associated with accelerated musculoskeletal aging, with the three largest associations being with right arm impedance (correlation=.049), right leg impedance (correlation=.044), and left arm impedance (correlation=.042). 53.3% of pulse wave biomarkers are associated with accelerated musculoskeletal aging, with the three largest associations being with heart rate (correlation=.041), central systolic blood pressure (correlation=.040) and brachial blood pressure (correlation=.040). 39.5% of brain MRI weighted means biomarkers are associated with accelerated musculoskeletal aging, with the three largest associations being with measurements in the left and right tract anterior thalamic radiations (correlations=.092-.089).

Conversely, the three biomarkers categories most associated with decelerated musculoskeletal aging are hand grip strength, cognitive symbol digit substitution and heel bone densitometry. Specifically, the two hand grip strength biomarkers are associated with decelerated musculoskeletal aging (right and left hand grip correlations are respectively .123 and .115). The two symbol digits substitutions (a cognitive test) biomarkers are associated with decelerated musculoskeletal aging (number of symbol digit matches made correctly: correlation=.058. Number of symbol digit matches attempted: correlation=.058). 83.3% of heel bone densitometry biomarkers are associated with decelerated musculoskeletal aging, with the three largest correlations being with speed of sound through heel (correlation=.063), heel bone mineral density (correlation=.062) and heel quantitative ultrasound index (correlation=.062).

### Clinical phenotypes associated with accelerated musculoskeletal aging

The three clinical phenotype categories most associated with accelerated musculoskeletal aging are breathing, claudication, and self-reported “general health”. Specifically, the two breathing phenotypes are associated with accelerated musculoskeletal aging (shortness of breath walking on level ground: correlation=.041, wheeze or whistling in the chest in the last year: correlation=.027). 53.8% of the claudication phenotypes are associated with accelerated musculoskeletal aging, with the three largest associations being leg pain on walking (correlation=.066), leg pain when walking uphill or hurrying (correlation=.061), and leg pain on walking: action taken (correlation=.057). 50.0% of general health phenotypes are associated with accelerated musculoskeletal aging, with the three largest associations being with overall health rating (correlation=.082), losing weight during the last year (correlation=.076), and long-standing illness, disability or infirmity (correlation=.075).

Conversely, the three clinical phenotype categories most associated with decelerated musculoskeletal aging are sexual factors (age first had sexual intercourse: correlation=.031), cancer screening (most recent bowel cancer screening: correlation=.029) and mouth health (no mouth/dental problem: correlation=.027).

### Diseases associated with accelerated musculoskeletal aging

The three disease categories most associated with accelerated musculoskeletal aging are musculoskeletal diseases, cardiovascular diseases and general disease state. Specifically, 7.7% of musculoskeletal diseases are associated with accelerated musculoskeletal aging, with the three largest associations being with knee arthrosis (correlation=.046), rheumatoid arthritis (correlation=.042), and arthrosis (correlation=.038). 6.5% of cardiovascular diseases are associated with accelerated musculoskeletal aging, with the three largest associations being with hypertension (correlation=.059), chronic ischaemic heart disease (correlation=.033), and angina pectoris (correlation=.031). 4.8% of general disease state variables are associated with accelerated musculoskeletal aging, with the three largest associations being with personal history of disease (correlation=.043), problems related to lifestyle (correlation=.038), and personal history of medical treatment (correlation=.036).

### Environmental variables associated with accelerated musculoskeletal aging

The three environmental variable categories most associated with accelerated musculoskeletal aging are smoking, sun exposure, and electronic device usage. Specifically, 37.5% of smoking variables are associated with accelerated musculoskeletal aging, with the three largest associations being with pack years adult smoking as proportion of lifespan exposed to smoking (correlation=.084), pack years of smoking (correlation=.080), and number of cigarettes currently smoked daily (correlation=.072). 25.0% of sun exposure variables are associated with accelerated musculoskeletal aging, with the three largest associations being with time spent outdoors in summer (correlation=.072), time spent outdoors in winter (correlation=.060), and facial aging: not knowing whether one looks younger or older than one’s chronological age (correlation=.052). One electronic devices variable is associated with accelerated musculoskeletal aging (using mobile phone on the left side: correlation=.027).

Conversely, the three environmental variable categories most associated with decelerated musculoskeletal aging are physical activity, smoking and medication. Specifically, 34.3% of physical activity variables are associated with decelerated musculoskeletal aging, with the three largest associations being with the frequency of strenuous sports in the last four weeks (correlation=.070), duration of strenuous sports (correlation=.069), and the frequency of other exercises in the last four weeks (correlation=.065). 25.0% of smoking variables are associated with decelerated musculoskeletal aging, with the three largest associations being time from waking to first cigarette (correlation=.073), age started smoking (correlation=.070), and smoking status: never smoked (correlation=.068). 13.5% of medication variables are associated with decelerated musculoskeletal aging, with the three largest associations being with not taking any medication for cholesterol, blood pressure or diabetes (correlation=.074), not taking any medication for pain relief, constipation or heartburn (correlation=.041), and not taking any vitamins or supplements (correlation=.030).

### Socioeconomic variables associated with accelerated musculoskeletal aging

The three socioeconomic variable categories most associated with accelerated musculoskeletal aging are socio-demographics (private healthcare: correlation=.034), education (no degree: correlation=.032) and household (renting accommodation from local authority, local council or housing association: correlation=.040).

Conversely, the three socioeconomic variable categories most associated with decelerated musculoskeletal aging are education, socio-demographics and employment. Specifically, 25.0% of education variables are associated with decelerated musculoskeletal aging, with the two associations being college or university degree (correlation=.054) and A/AS levels or equivalent (correlation=.033). One sociodemographic variable is associated with decelerated musculoskeletal aging: not receiving any attendance/disability/mobility allowance (correlation=.052). 13.0% of employment variables are associated with decelerated musculoskeletal aging, with the three largest associations being with the length of working week for main job (correlation=.055), current employment status: in paid employment or self-employed (correlation=.053), and frequency of travelling from home to job workplace (correlation=.030).

### Predicting accelerated aging from biomarkers, clinical phenotypes, diseases, environmental variables and socioeconomic variables

We predicted accelerated musculoskeletal aging using variables from the different X-datasets categories (biomarkers, clinical phenotypes, diseases, environmental variables and socioeconomic variables). Specifically we built a model using the variables from each of their respective subcategories (e.g blood pressure biomarkers), and found that no dataset could explain more than 5% of the variance in accelerated musculoskeletal aging.

### Phenotypic, genetic and environmental correlation between the different musculoskeletal aging dimensions

#### Phenotypic correlation between aging dimensions

To determine whether the different components of the musculoskeletal systems of a given individual age at the same rate, we computed the Pearson correlation between accelerated aging (predicted age - chronological age) in each musculoskeletal aging dimension (Fig. S11 and Figure 6 - upper left matrix). We found an average correlation of .272±.130, based on the ten pairwise correlations. The correlation increases as the aging dimensions become more similar. Specifically, we found that the different X-ray-based dimensions are .351±.072 correlated on average (six pairwise correlations), and the scalar-based subdimensions (anthropometry, impedance, heel bone densitometry and hand grip strength) are .345±.092 correlated on average (six pairwise correlations). Sagittal view-based and coronal view-based accelerated spine aging are .487 correlated, “flesh view”-based and “figure view”-based accelerated full body aging are .543 correlated, and “figure view”-based and “skeleton view”-based accelerated full body aging are .714±.004 correlated.

**Figure 6:**
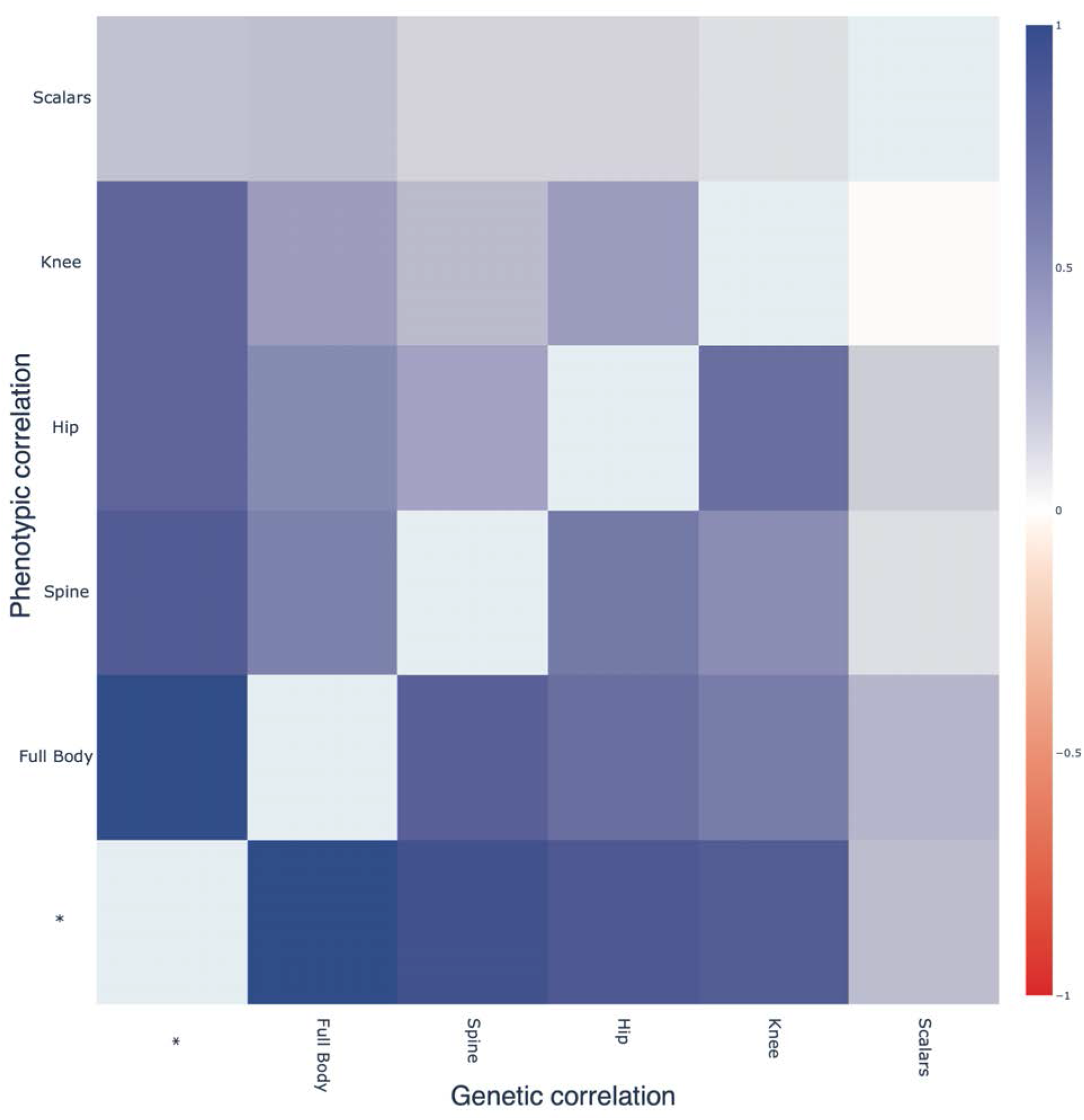
Phenotypic and genetic correlation between accelerated aging in the different musculoskeletal aging dimensions. Upper left triangle: Pearson correlation between accelerated aging for each pair of musculoskeletal dimensions. Lower right triangle: GWAS-based genetic correlation between accelerated aging for each pair of musculoskeletal dimensions.

#### Genetic correlation between aging dimensions

Images-based musculoskeletal aging dimensions (full body, spine, hip and knee) are genetically .494±.064 correlated (based on six genetic correlations), whereas the scalar features-based dimension (anthropometry, impedance, heel bone densitometry and hand grip strength) is only .126±.098 correlated (based on four correlations) with the images-based musculoskeletal dimensions. (Figure 6 - lower right matrix)

#### Correlation between musculoskeletal aging dimensions in terms of associations with biomarkers, clinical phenotypes, diseases

We compared associations with non-genetic variables across the different musculoskeletal aging dimensions to understand if X-variables associated with accelerated aging in one musculoskeletal dimension are also associated with accelerated aging in another musculoskeletal dimension. For example, in terms of environmental variables, are the diets that protect against spine aging the same as the diets that protect against knee aging?

We found that the average correlation between musculoskeletal aging dimensions is .461±.325 in terms of biomarkers, .784±.120 in terms of associated clinical phenotypes, .354±.278 in terms of diseases, .850±.187 in terms of family history, .608±.190 in terms of environmental variables and .608+-.190 in terms of socioeconomic variables (Fig. S12). To compare two specific musculoskeletal dimensions, please refer to https://www.multidimensionality-of-aging.net/correlation_between_aging_dimensions/xwas_univariate - tab “Summary”, where they can be interactively explored. For the sake of the example, we provide the correlations between accelerated hip aging and accelerated knee aging in terms of the different X-associations in Fig. S13.

## Discussion

### We built the most accurate musculoskeletal age predictor to date

We built what is, to the best of our knowledge, the most accurate musculoskeletal system-based chronological age predictor (RMSE=2.65±0.01 years; R-Squared=87.6±0.1%). Specifically, we built the first spine X-ray-based chronological age predictor (R^2^=74.6±0.2%; RMSE=3.81±0.01 years) and we outperformed the best chronological age predictors in the literature for full body X-rays (R^2^=85.7±0.1%; RMSE=2.85±0.01 years vs. R^2^=83%), hip X-rays (R^2^=69.0±0.3%; RMSE=4.20±0.02 years) and adult knee X-rays, (R^2^=69.0±0.3%; RMSE=4.20±0.02 years). A summary of the comparison between our models and the models reported in the literature can be found in Table S1. We describe and discuss these comparisons more in detail in the Supplementary.

### Anatomical features driving age prediction

The attention maps of the models trained on knee X-ray images highlighted the joint, possibly relying on osteoarthritis ^32, 33^. Similarly, the attention maps of the models trained on hip X-ray images highlighted both the joint and the proximal femur, which can possibly be explained by osteoporosis ^34, 35^ and changes in the shapes of the femoral neck ^36^ and medullary cavity ^37^ with age. The attention maps of the models trained on spine X-ray images highlighted different vertebrae, which can be explained by osteoporosis and degenerative changes ^38^. The lumbar spine was particularly highlighted and is known to suffer from disc degeneration, among other changes^7^. Finally, the attention maps of the models trained on full body X-ray images highlighted the neck, the upper torso, the hips and the knees. The highlighting of the neck can be possibly explained by the fact that the neck tilt and the cervical angle tend to increase with age ^39^ and the highlighting of the upper torso can be explained by increased osteoporosis and thoracic kyphosis with age, as well as changes in the ribs shapes and angles ^40, 41^.

With age, the body composition changes, as muscle mass decreases whereas fat mass increases ^42–44^, which explains why the models trained on scalar datasets selected hand grip strength, waist circumference, weight, whole-body impedance and body mass index as important age predictors.

### We identified genetic factors associated with accelerated musculoskeletal aging

We found that accelerated musculoskeletal aging is 34.9±1.8% GWAS-heritable, with the GWAS-heritability of its different dimensions ranging from 32.9±1.7% for spine aging to 22.1±0.1 for scalar biomarkers-based (anthropometry, impedance, heel bone densitometry, hand grip strength) aging. We explain this difference in heritability, in part, by the difference in chronological age prediction performances for the different dimensions (Figure 2). This suggests that improving the chronological age predictors would lead to an increase in the estimated GWAS-heritability of the associated accelerated musculoskeletal aging dimensions.

Among the GWAS peaks we identified and investigated, approximately half include a gene whose function has a clear connection to musculoskeletal health (e.g CCDC91, associated with ossification of the posterior longitudinal ligament of the spine and diffuse idiopathic skeletal hyperostosis), providing insights about the way genetics can influence the aging rates of the different musculoskeletal dimensions. The GWAS highlighted other genes linked to the cardiovascular health (e.g FGFR3P1, involved in vascular endothelial growth factor signaling), cancer (e.g HTRA1, involved in cell growth regulation; FAM3C, linked to pancreatic cancer), skin health (e.g AAGAB, linked to palmoplantar keratoderma), blood count (e.g SOCS2, involved in cytokine signaling and linked to polycythemia) and kidney health (e.g PKD2L1, linked to polycystic kidney disease). These findings suggest that accelerated musculoskeletal aging is associated with poor health in other organ systems. In particular, we found that the most significant SNP associated with accelerated knee aging lies in the WNT16 gene, which is involved in signaling and is linked to aging in different organ systems ^45–48^, such as the brain ^49^, the heart ^50^ and the intestines ^51^. We explore the hypothesis of multidimensional aging more in detail in a different paper^52^. Genes associated with accelerated musculoskeletal aging represent potentially promising target candidates for rejuvenating therapies.

### We identified non-genetic factors associated with accelerated musculoskeletal aging

Accelerated musculoskeletal aging is associated with musculoskeletal biomarkers such as anthropometry, impedance and hand grip strength, as well as non musculoskeletal biomarkers such as cardiovascular biomarkers (e.g blood pressure, arterial stiffness, ECG, heart function), brain biomarkers (e.g cognitive function, MRI-features), blood biomarkers (e.g biochemistry, blood count) and hearing biomarkers. We observed a similar pattern in terms of association with clinical phenotypes and diseases.

Accelerated musculoskeletal aging is associated not only with musculoskeletal phenotypes (claudication, joint pain) and diseases (e.g knee arthrosis and rheumatoid arthritis), but also with non-musculoskeletal phenotypes (e.g chest pain, mental health, mouth health) and diseases (e.g cardiovascular, respiratory, mental, metabolic), suggesting that accelerated musculoskeletal aging is correlated with accelerated aging in other organ systems. More generally, musculoskeletal aging is associated with general health (e.g overall health rating, weight loss during the last year, long-standing illness, disability or infirmity, general history of disease/medical treatment, unhealthy lifestyle), and is observable to some degree, as it is correlated with looking older than one’s age. These observations suggest that musculoskeletal aging is affected by a general aging factor that similarly affects the aging of other organ systems. However, again, while correlations were statistically significant, their association sizes were modest to negligible. We further explore the connection between musculoskeletal aging and aging in other organ systems in a different paper ^52^.

We found that height at age ten and accelerated musculoskeletal aging at adulthood are positively correlated. A possible explanation is that taller individuals suffer from increased stress on their joints, leading to premature musculoskeletal aging.

We identified lifestyle factors associated with accelerated musculoskeletal aging (e.g smoking) and decelerated musculoskeletal aging (e.g physical activity), which is coherent with the literature^53, 54^ and credits the potential of lifestyle interventions to slow or reverse musculoskeletal aging. UKB being an observational dataset, we however cannot claim to have proven a causal link here.

We found that accelerated musculoskeletal aging to be negatively correlated to socioeconomic status (e.g income, education), reflecting the literature. In the US, the richest 1% males live on average 14.6±0.2 years longer than the poorest 1% males, and the richest 1% females live on average 10.1±0.2 years longer than the poorest 1% females ^55^. These differences are likely due in part to better healthcare access and health literacy ^56^.

### The different musculoskeletal components of a single individual can age at different rates

We found that, on average (based on ten pairwise correlations), the different musculoskeletal aging dimensions are phenotypically .272±.130 correlated and genetically .347±.208 correlated. This correlation increases when the musculoskeletal dimensions considered are more similar, for example when comparing X-ray-based musculoskeletal dimensions. While these correlations might still seem low, one should keep in mind that the low values can be in part explained by the variance of the age predictors and their imperfect predictions. For example, two convolutional neural network architectures (InceptionV3 and InceptionResNetV2) trained on the exact same dataset (full body “Figure” view X-ray images) yielded accelerated aging definitions that are only .717±.003 correlated. Therefore, the perhaps surprisingly low phenotypic correlation between hip aging and knee aging (.351±.004) can be reinterpreted as being approximately half the correlation observed between two biologically identical datasets.

In contrast to the low phenotypic and genetic correlations between musculoskeletal dimensions, we found that the different dimensions tend to be similarly associated with some of the biomarkers, clinical phenotypes, diseases, environmental and socioeconomic variables categories. For example, smoking is associated with accelerated aging in all musculoskeletal dimensions (average correlation between two musculoskeletal dimensions in terms of association between accelerated aging and smoking variables = .825±.153). In other words, exposures that lead to accelerated aging of a specific joint are likely to be associated with accelerated aging of other joints, as well. While the association of these non-genetic variables tend to be shared across musculoskeletal aging dimensions, it should be noted that each non-genetic variable category explained less than 5% of the variance in accelerated musculoskeletal aging.

### Limitations

Some of our predictors predicted chronological age with high accuracy (e.g all encompassing ensemble model: RMSE=2.65±0.04), and it has been suggested that, as chronological age prediction accuracy increases, the associated biological age predictors lose their clinical significance ^57, 58^. A model that perfectly predicts chronological age only outputs chronological age, not biological age.

In terms of associations between accelerated aging and non-genetic variables such as environmental exposures, UKB is an observational study, which means that observing these correlations does not allow us to infer causality. Each correlation could potentially be explained by direct causality (e.g not exercising leads to accelerated musculoskeletal aging), reverse causality (e.g poor musculoskeletal health limits physical activity) or confounding factors (e.g exposure to a chemical could lead to both decreased musculoskeletal function and lung function, the later leading to decreased physical activity).

### Utility of musculoskeletal age predictors

In conclusion, our predictors can be used to monitor the aging process of different musculoskeletal components of a single individual and suggest potential lifestyle and therapeutic interventions to slow musculoskeletal aging. Finally, we hypothesize that musculoskeletal system-specific biological age predictors could be used in clinical trials to assess the effect of rejuvenating therapies ^59^ for musculoskeletal aging. Because aging is largely multidimensional ^52, 60^, general biological age predictors, such as the DNA methylation clock ^61–63^, might fail to measure the efficiency of rejuvenating drug candidates on this specific facet of aging.

## Methods

### Data and materials availability

We used the UK Biobank (project ID: 52887). The code can be found at https://github.com/Deep-Learning-and-Aging. The results can be interactively and extensively explored at https://www.multidimensionality-of-aging.net/. We will make the biological age phenotypes available through UK Biobank upon publication. The GWAS results can be found at https://www.dropbox.com/s/59e9ojl3wu8qie9/Multidimensionality_of_aging-GWAS_results.zip?dl=0.

### Software

Our code can be found at https://github.com/Deep-Learning-and-Aging. For the genetics analysis, we used the BOLT-LMM ^64, 65^ and BOLT-REML ^66^ softwares. We coded the parallel submission of the jobs in Bash ^67^.

### Cohort Dataset: Participants of the UK Biobank

We leveraged the UK Biobank^31^ cohort (project ID: 52887). The UKB cohort consists of data originating from a large biobank collected from 502,211 de-identified participants in the United Kingdom that were aged between 37 years and 74 years at enrollment (starting in 2006). Out of these participants, 46,572 had X-ray images taken. The Harvard internal review board (IRB) deemed the research as non-human subjects research (IRB: IRB16-2145).

### Data types and Preprocessing

The data preprocessing step is different for the different data modalities: demographic variables, scalar predictors and images. We define scalar predictors as predictors whose information can be encoded in a single number, such as height, as opposed to data with a higher number of dimensions such as images (two dimensions, which are the height and the width of the image).

#### Demographic variables

First, we removed out the UKB samples for which age or sex was missing. For sex, we used the genetic sex when available, and the self-reported sex when genetic sex was not available. We computed age as the difference between the date when the patient attended the assessment center and the year and month of birth of the patient to estimate the patient’s age with greater precision. We one-hot encoded ethnicity.

#### Scalar biomarkers: anthropometry, impedance, heel bone densitometry and hand grip strength

We define scalar data as a variable that is encoded as a single number, such as height or right arm impedance, as opposed to data with a higher number of dimensions, such as images. The complete list of scalar biomarkers can be found in Table S5 under “Musculoskeletal”. We did not preprocess the scalar data, aside from the normalization that is described under cross-validation further below.

#### X-ray images

The UKB contains skeletal Dual-energy X-ray absorptiometry (DXA) images from the spine, hips, knees and full body (field 20158, 42,287 images for 41,212 participants), saved as DICOM files.

For the spine, images were collected for both the sagittal plane and the coronal plane. Not all images had the same dimensions, so we resized all of them to the median size (1513*684 pixels for sagittal, 724*720 pixels for coronal) (Supplementary Figure 62).

For the hip and knee images, we took the left symmetry for the left side images. The images had different dimensions between the participants, so we resized all the images to respectively 626*680 (hips) and 851*700 (knees). A sample for hip and knee images can respectively be found in Supplementary Figure 63 and Supplementary Figure 64.

Two full body images are made available by UKB. One displays the skeleton of the participant, surrounded by the shape of their body. We refer to this image as “Figure”. The other barely displays the bone structure but makes the other tissues more visible. We refer to this image as “Flesh”. From these two images, we generated two more images for each participant. (1) An image for which we removed the shape of the body surrounding the skeleton, which we named “Skeleton”. To isolate the skeleton from the rest of the image, we used the following algorithm. For each pixel at the border of the image, we computed the position of the first bone pixel encountered in every direction. All the pixels between the border pixel and the bone pixel were set to black. We defined the bone pixels to be pixels with a value larger than or equal to 70 (white color) and smaller than 250 (to exclude non-bone bright artefact pixels). (2) An image for which we superposed the three first images (Figure, Flesh and Skeleton) to generate a three-layer RGB image that we named “Mixed”. We resized these four images to 811*272 pixels, the median image size, for all participants (Supplementary Figure 61).

We rescaled the values of the pixels to be between 0 and 1 instead of between 0 and 255 to mitigate the explosion of gradients in the neural networks (see Methods-Algorithms). Starting from grayscale images, we saved them as RGB by duplicating the values three times. As explained in detail further below, we used transfer learning to analyze the images. In the context of transfer learning, the size of the architecture of the model is affected by the dimension of the input. Large images contain more information but require larger deep learning architectures that take longer to train. To resolve prohibitory long training times, we resized the images so that the total number of pixels for each channel would be below 100,000.

#### Data augmentation

To prevent overfitting and increase our sample size during the training we used data augmentation^68^ on the images. Each image was randomly shifted vertically and horizontally, as well as rotated and zoomed. We chose the hyperparameters for these transformations’ distributions to represent the variations we observed between the images in the initial dataset. For example, for the hip and knee images, we observed similar variation between images in the vertical and the horizontal direction, so both the random vertical and horizontal shifts were sampled from the [-10%; +10%] uniform distribution. A summary of the hyperparameter values for the transformations’ distributions can be found in Supplementary Table 37.

The data augmentation process is dynamically performed during the training. Augmented images are not generated in advance. Instead, each image is randomly augmented before being fed to the neural network for each epoch during the training.

### Machine learning algorithms

For scalar datasets, we used elastic nets, gradient boosted machines [GBMs] and fully connected neural networks. For images we used two-dimensional convolutional neural networks.

#### Scalar data

We used three different algorithms to predict age from scalar data (non-dimensional variables, such as laboratory values). Elastic Nets [EN] (a regularized linear regression that represents a compromise between ridge regularization and LASSO regularization), Gradient Boosted Machines [GBM] (LightGBM implementation ^69^), and Neural Networks [NN]. The choice of these three algorithms represents a compromise between interpretability and performance. Linear regressions and their regularized forms (LASSO ^70^, ridge ^71^, elastic net ^72^) are highly interpretable using the regression coefficients but are poorly suited to leverage non-linear relationships or interactions between the features and therefore tend to underperform compared to the other algorithms. In contrast, neural networks ^73, 74^ are complex models, which are designed to capture non-linear relationships and interactions between the variables. However, tools to interpret them are limited ^75^ so they are closer to a “black box”. Tree-based methods such as random forests ^76^, gradient boosted machines ^77^ or XGBoost ^78^ represent a compromise between linear regressions and neural networks in terms of interpretability. They tend to perform similarly to neural networks when limited data is available, and the feature importances can still be used to identify which predictors played an important role in generating the predictions. However, unlike linear regression, feature importances are always non-negative values, so one cannot interpret whether a predictor is associated with older or younger age. We also performed preliminary analyses with other tree-based algorithms, such as random forests ^76^, vanilla gradient boosted machines ^77^ and XGBoost ^78^. We found that they performed similarly to LightGBM, so we only used this last algorithm as a representative for tree-based algorithms in our final calculations.

#### X-ray images

##### Convolutional Neural Networks Architectures

We used transfer learning ^79–81^ to leverage two different convolutional neural networks ^82^ [CNN] architectures pre-trained on the ImageNet dataset ^83–85^ and made available through the python Keras library ^86^: InceptionV3 ^87^ and InceptionResNetV2 ^88^. We considered other architectures such as VGG16 ^89^, VGG19 ^89^ and EfficientNetB7 ^90^, but found that they performed poorly and inconsistently on our datasets during our preliminary analysis and we therefore did not train them in the final pipeline. For each architecture, we removed the top layers initially used to predict the 1,000 different ImageNet images categories. We refer to this truncated model as the “base CNN architecture”.

We added to the base CNN architecture what we refer to as a “side neural network”. A side neural network is a single fully connected layer of 16 nodes, taking the sex and the ethnicity variables of the participant as input. The output of this small side neural network was concatenated to the output of the base CNN architecture described above. This architecture allowed the model to consider the features extracted by the base CNN architecture in the context of the sex and ethnicity variables. For example, the presence of the same anatomical feature can be interpreted by the algorithm differently for a male and for a female. We added several sequential fully connected dense layers after the concatenation of the outputs of the CNN architecture and the side neural architecture. The number and size of these layers were set as hyperparameters. We used ReLU ^91^ as the activation function for the dense layers we added, and we regularized them with a combination of weight decay ^92, 93^ and dropout ^94^, both of which were also set as hyperparameters. Finally, we added a dense layer with a single node and linear activation to predict age.

##### Compiler

The compiler uses gradient descent ^95, 96^ to train the model. We treated the gradient descent optimizer, the initial learning rate and the batch size as hyperparameters. We used mean squared error [MSE] as the loss function, root mean squared error [RMSE] as the metric and we clipped the norm of the gradient so that it could not be higher than 1.0 ^97^.

We defined an epoch to be 32,768 images. If the training loss did not decrease for seven consecutive epochs, the learning rate was divided by two. This is theoretically redundant with the features of optimizers such as Adam, but we found that enforcing this manual decrease of the learning rate was sometimes beneficial. During training, after each image has been seen once by the model, the order of the images is shuffled. At the end of each epoch, if the validation performance improved, the model’s weights were saved.

We defined convergence as the absence of improvement on the validation loss for 15 consecutive epochs. This strategy is called early stopping ^98^ and is a form of regularization. We requested the GPUs on the supercomputer for ten hours. If a model did not converge within this time and improved its performance at least once during the ten hours period, another GPU was later requested to reiterate the training, starting from the model’s last best weights.

### Training, tuning and predictions

We split the entire dataset into ten data folds. We then tuned the models built on scalar data and the models built on images using two different pipelines. For scalar data-based models, we performed a nested-cross validation. For images-based models, we manually tuned some of the hyperparameters before performing a simple cross-validation. We describe the splitting of the data into different folds and the tuning procedures in greater detail in the Supplementary.

### Interpretability of the machine learning predictions

To interpret the models, we used the regression coefficients for the elastic nets, the feature importances for the GBMs, a permutation test for the fully connected neural networks, and attention maps (saliency and Grad-RAM) for the convolutional neural networks (Supplementary Methods).

### Ensembling to improve prediction and define aging dimensions

We built a three-level hierarchy of ensemble models to improve prediction accuracies. At the lowest level, we combined the predictions from different algorithms on the same aging subdimension. For example, we combined the predictions generated by the elastic net, the gradient boosted machine and the neural network from the anthropometric scalar biomarkers. At the second level, we combined the predictions from different subdimensions of a unique musculoskeletal dimension. For example, we combined the scalar subdimensions “Anthropometry”, “Impedance”, “Heel bone densitometry” and “Hand grip strength” into an ensemble prediction. Finally, at the highest level, we combined the predictions from the five musculoskeletal aging dimensions (full body, spine, hip, knee, scalar biomarkers) into a general musculoskeletal age prediction. The ensemble models from the lower levels are hierarchically used as components of the ensemble models of the higher models. For example, the ensemble model built by combining the algorithms trained on anthropometric variables is leveraged when building the general musculoskeletal aging ensemble model.

We built each ensemble model separately on each of the ten data folds. For example, to build the ensemble model on the testing predictions of the data fold #1, we trained and tuned an elastic net on the validation predictions from the data fold #0 using a 10-folds inner cross-validation, as the validation predictions on fold #0 and the testing predictions on fold #1 are generated by the same model (see Methods - Training, tuning and predictions - Images - Scalar data - Nested cross-validation; Methods - Training, tuning and predictions - Images - Cross-validation). We used the same hyperparameters space and Bayesian hyperparameters optimization method as we did for the inner cross-validation we performed during the tuning of the non-ensemble models.

To summarize, the testing ensemble predictions are computed by concatenating the testing predictions generated by ten different elastic nets, each of which was trained and tuned using a 10-folds inner cross-validation on one validation data fold (10% of the full dataset) and tested on one testing fold. This is different from the inner-cross validation performed when training the non-ensemble models, which was performed on the “training+validation” data folds, so on 9 data folds (90% of the dataset).

### Evaluating the performance of models

We evaluated the performance of the models using two different metrics: R-Squared [R^2^] and root mean squared error [RMSE]. We computed a confidence interval on the performance metrics in two different ways. First, we computed the standard deviation between the different data folds. The test predictions on each of the ten data folds are generated by ten different models, so this measure of standard deviation captures both model variability and the variability in prediction accuracy between samples. Second, we computed the standard deviation by bootstrapping the computation of the performance metrics 1,000 times. This second measure of variation does not capture model variability but evaluates the variance in the prediction accuracy between samples.

### Musculoskeletal age definition

We defined the biological age of participants as the prediction generated by the model corresponding to musculoskeletal dimension or subdimension, after correcting for the bias in the residuals.

We indeed observed a bias in the residuals. For each model, participants on the older end of the chronological age distribution tend to be predicted younger than they are. Symmetrically, participants on the younger end of the chronological age distribution tend to be predicted older than they are. This bias does not seem to be biologically driven. Rather it seems to be statistically driven, as the same 60-year-old individual will tend to be predicted younger in a cohort with an age range of 60-80 years, and to be predicted older in a cohort with an age range of 60-80. We ran a linear regression on the residuals as a function of age for each model and used it to correct each prediction for this statistical bias.

After defining biological age as the corrected prediction, we defined accelerated aging as the corrected residuals. For example, a 60-year-old whose spine X-ray data predicted an age of 70 years old after correction for the bias in the residuals is estimated to have a spine age of 70 years, and an accelerated spine aging of ten years.

It is important to understand that this step of correction of the predictions and the residuals takes place after the evaluation of the performance of the models but precedes the analysis of the musculoskeletal ages properties.

### Genome-wide association of accelerated musculoskeletal aging

The UKB contains genome-wide genetic data for 488,251 of the 502,492 participants^99^ under the hg19/GRCh37 build.

We used the average accelerated aging value over the different samples collected over time (see Supplementary - Models ensembling - Generating average predictions for each participant). Next, we performed genome wide association studies [GWASs] to identify single-nucleotide polymorphisms [SNPs] associated with accelerated aging in each musculoskeletal dimension using BOLT-LMM ^64, 65^ and estimated the the SNP-based heritability for each of our biological age phenotypes, and we computed the genetic pairwise correlations between dimensions using BOLT-REML ^66^. We used the v3 imputed genetic data to increase the power of the GWAS, and we corrected all of them for the following covariates: age, sex, ethnicity, the assessment center that the participant attended when their DNA was collected, and the 20 genetic principal components precomputed by the UKB. We used the linkage disequilibrium [LD] scores from the 1,000 Human Genomes Project ^100^. To avoid population stratification, we performed our GWAS on individuals with White ethnicity.

#### Identification of SNPs associated with accelerated aging

We identified the SNPs associated with accelerated musculoskeletal aging dimensions and subdimensions using the BOLT-LMM ^64, 65^ software (p-value of 5e-8). The sample size for the genotyping of the X chromosome is one thousand samples smaller than for the autosomal chromosomes. We therefore performed two GWASs for each aging dimension. (1) excluding the X chromosome, to leverage the full autosomal sample size when identifying the SNPs on the autosome. (2) including the X chromosome, to identify the SNPs on this sex chromosome. We then concatenated the results from the two GWASs to cover the entire genome, at the exception of the Y chromosome.

We plotted the results using a Manhattan plot and a volcano plot. We used the bioinfokit ^101^ python package to generate the Manhattan plots. We generated quantile-quantile plots [Q-Q plots] to estimate the p-value inflation as well.

#### Heritability and genetic correlation

We estimated the heritability of the accelerated aging dimensions using the BOLT-REML ^66^ software. We included the X chromosome in the analysis and corrected for the same covariates as we did for the GWAS. Using the same software and parameters, we computed the genetic correlations between accelerated aging in the different musculoskeletal dimensions.

We annotated the significant SNPs with their matching genes using the following four steps pipeline. (1) We annotated the SNPs based on the rs number using SNPnexus ^102–106^. When the SNP was between two genes, we annotated it with the nearest gene. (2) We used SNPnexus to annotate the SNPs that did not match during the first step, this time using their genomic coordinates. After these two first steps, 30 out of the 9,697 significant SNPs did not find a match. (3) We annotated these SNPs using LocusZoom ^107^. Unlike SNPnexus, LocusZoom does not provide the gene types, so we filled this information with GeneCards ^108^. After this third step, four genes were not matched. (4) We used RCSB Protein Data Bank ^109^ to annotate three of the four missing genes.

### Non-genetic correlates of accelerated aging

We identified non-genetically measured (i.e factors not measured on a GWAS array) correlates of each aging dimension, which we classified in six categories: biomarkers, clinical phenotypes, diseases, family history, environmental, and socioeconomic variables. We refer to the union of these association analyses as an X-Wide Association Study [XWAS]. (1) We define as biomarkers the scalar variables measured on the participant, which we initially leveraged to predict age (e.g. blood pressure, Table S5). (2) We define clinical phenotypes as other biological factors not directly measured on the participant, but instead collected by the questionnaire, and which we did not use to predict chronological age. For example, one of the clinical phenotypes categories is eyesight, which contains variables such as “wears glasses or contact lenses”, which is different from the direct refractive error measurements performed on the patients, which are considered “biomarkers” (Table S8). (3) Diseases include the different medical diagnoses categories listed by UKB (Table S11). (4) Family history variables include illnesses of family members (Table S14). (5) Environmental variables include alcohol, diet, electronic devices, medication, sun exposure, early life factors, medication, sun exposure, sleep, smoking, and physical activity variables collected from the questionnaire (Table S17). (6) Socioeconomic variables include education, employment, household, social support and other sociodemographics (Table S20). We provide information about the preprocessing of the XWAS in the Supplementary Methods.

## Data Availability

https://github.com/alanlegoallec/Multidimensionality_of_Aging

https://www.dropbox.com/s/59e9ojl3wu8qie9/Multidimensionality_of_aging-GWAS_results.zip?dl=0

## Author Contributions

**Alan Le Goallec:** (1) Designed the project. (2) Supervised the project. (3) Predicted chronological age from images. (4) Computed the attention maps for the images. (5) Ensembled the models, evaluated their performance, computed biological ages and estimated the correlation structure between the musculoskeletal aging dimensions. (6) Performed the genome wide association studies. (5) Designed the website. (6) Wrote the manuscript.

**Samuel Diai:** (1) Predicted chronological age from scalar features. (2) Coded the algorithm to obtain balanced data folds across the different datasets. (3) Wrote the python class to build an ensemble model using a cross-validated elastic net. (4) Performed the X-wide association study. (5) Implemented a first version of the website https://www.multidimensionality-of-aging.net/.

**Sasha Collin:** (1) Preprocessed the X-ray images.

**Théo Vincent:** (1) Website data engineer. (2) Implemented a second version of the website https://www.multidimensionality-of-aging.net/.

**Chirag J. Patel:** (1) Supervised the project. (2) Edited the manuscript. (3) Provided funding.

## Acknowledgments

We would like to thank Raffaele Potami from Harvard Medical School research computing group for helping us utilize O2’s computing resources. We also want to acknowledge UK Biobank for providing us with access to the data they collected. The UK Biobank project number is 52887. We thank HMS RC for computing support.

## Funding

NIEHS R00 ES023504

NIEHS R21 ES25052.

NIAID R01 AI127250

NSF 163870

MassCATS, Massachusetts Life Science Center

Sanofi

The funders had no role in the study design or drafting of the manuscript(s).

